# Fuzzy logic use in classification of the severity of diabetic retinopathy

**DOI:** 10.1101/2020.05.11.20098756

**Authors:** Luís Jesuino de Oliveira Andrade, Caroline Santos França, Rafael Andrade, Alcina Maria Vinhaes Bittencourt, Gabriela Correia Matos de Oliveira

## Abstract

**Purpose:** Employ fuzzy logic to auxiliary in identification and diagnosis the gravity of diabetic retinopathy (DR).

**Methods:** A cross-sectional study was performed, being assessed 100 diabetes mellitus patients with DR. The following ultrasound findings were measured employing a semi-quantitative punctuation method: vitreous hemorrhage, posterior vitreous detachment, epiretinal fibrosis, retinal detachment. The fundus photography (FP) aspects evaluated for diagnosis of DR were at least four or more microaneurysms with or without hard or soft exudates, and neovascularization, graded using the Early Treatment of Diabetic Retinopathy Scale. With the combination between ultrasound punctuation and FP aspects through fuzzy logic, a classification for DR has been built.

**Results:** Microaneurysms were the findings which presented the better interaction with the DR severity on ultrasound, while the hard exudates showed the minors estimation errors when compared to soft exudates. A classification for DR was suggested based on the 95% confidence interval of number of microaneurysms: mild group (< 24.6); moderately mild (24.6 - 48.0); moderate (48.1 - 64.5); moderately severe (64.6 - 77.0); severe (77.1 - 92.7); and very severe (> 92.7).

**Conclusion:** By the fuzzy logic, a DR classification was constructed supported on number of microaneurysms measurement with a simple practical application.

## Introduction

Diabetes mellitus (DM) is a public health problem, and diabetic retinopathy (DR) is a complication of DM and its detection is fundamental for prevention of blindness. The DR is neurovascular disease of retina which generates visual impairment and blindness in working-age diabetic patients. Study has evaluated that circa 93 million individuals must have DR, and approximately 28 million individuals may have DR in advanced stages^(1)^. The pathophysiology of DR is heterogeneous and includes inflammation and oxidative stress, having as a final result microvascular and neuronal disorder^(2)^. The microvascular complications of DR include microaneurysms, vascular occlusion, retinal neovascularization, retinal edema, vitreous hemorrhage, fibrovascular proliferation leading to retinal detachment, which is clinically represented by two stages, the early stage that called non-proliferative DR, and a more advanced stage denominated of proliferative DR^(3)^.

Although recent advances of ophthalmologic examination techniques, analyze of retinal changes DR secondary frequently features a diagnostic defiance. Usually, the diagnostic of DR mainly relies on fundus photography (FP) obtained by mydriatic or nonmydriatic fundus cameras that will be diagnosed by ophthalmy specialist. Ocular sonography is useful in detecting of DR, because the eye can be rated dynamically during ocular movements, principally in eyes with opaque media, besides being superior to computed tomography and magnetic resonance imaging in diagnosis of DR^(4)^.

Fuzzy logic performs a significant function in medicine, because the difficulties of medicine exercise become usual quantitative approaches of analysis inadequate. Fuzzy logic goes back to 1965, and it deals with common sense reasoning, in other words, with estimated logic. Fuzzy logic (‘almost certain’/‘very unlikely’), in contrast to classical propositional logic (true/false), is a multi-valued logic acquired starting fuzzy group theory agreement with the human argument^(5)^. Fuzzy logic is mathematical tool to deal with the inaccuracy and doubt human being, and has been applied in countless accost to model diagnosis. Thus, to analyze and diagnose early the diabetic retinopathy is one of the examples of advantage of using fuzzy logic.

The DR diagnosis is one of the essential steps of clinical conduct execution that in general is followed by a phase of insecurity and doubts. Hence, the aim of this study was outline fuzzy logic to auxiliary in identify and diagnosis gravity of DR.

## Methods

In this study we propose Fuzzy logic use for severity classification of DR through a cross-sectional study, being assessed 100 FP and ultrasonography of DM patients with DR.

The ocular ultrasonography was used to show secondary complications signals and define the Proliferative Diabetic Retinopathy stage, and correlate with FP.

### Ultrasound evaluation

Ultrasonography evaluation was fulfilled by a single physician with gray-scale sonographic equipment (Toshiba Aplio XG - Tokio, Japan) using a 10-12MHz linear multifrequency probe. The exams were interpreted by B-mode by transverse and longitudinal section of the four major eye quadrants, and following ultrasound findings were measured employing a semi-quantitative punctuation method: vitreous hemorrhage, posterior vitreous detachment, epiretinal fibrosis, and retinal detachment. The following values were given to the score of ultrasound ocular abnormality: 0= no retinal abnormality; 1= vitreous hemorrhage; 2= posterior vitreous detachment; 3= epiretinal fibrosis; 4= retinal detachment.

### Fundus photography evaluation

The gold standard photography method for diagnosis of DR is stereoscopic color FP in 7 standard fields (30°) as established by Early Treatment Diabetic Retinopathy Study (ETDRS) group^(6)^. The ETDRS severity level in the worse eye: minimal nonproliferative retinopathy, mild nonproliferative retinopathy, moderate nonproliferative retinopathy, severe nonproliferative retinopathy, and proliferative retinopathy. The FP evaluation was fulfilled by a single ophthalmy specialist with Canon CX-1 Digital Retinal Camera equipment. The FP aspects evaluated for diagnosis of DR were at least four or more microaneurysms with or without hard or soft exudates, and neovascularization. The following values were given to the score of FP abnormality: 0= no retinal abnormality; 1= four or more microaneurysms; 2= four or more microaneurysms without hard or soft exudates; 3= four or more microaneurysms with hard or soft exudates; 4= neovascularization.

### Study design and data analysis

With the combination between ultrasound punctuation and FP aspects through fuzzy logic, a classification for DR has been built.

The level of statistical significance was set at p < 0.05. Prism version 6.0 software free was used for data analysis (GraphPad Software Inc., La Jolla, CA, USA).

A DR classification was performed by the sharing among ultrasonography scores and FP scores through fuzzy logic. The fuzzy edge detection technique was grounded in a fuzzy inference system (Figure 1). The secondary alterations to DR were identified using the fuzzy k means, and a pre-processing was performed by histogram equalization.

**Figure 1.**
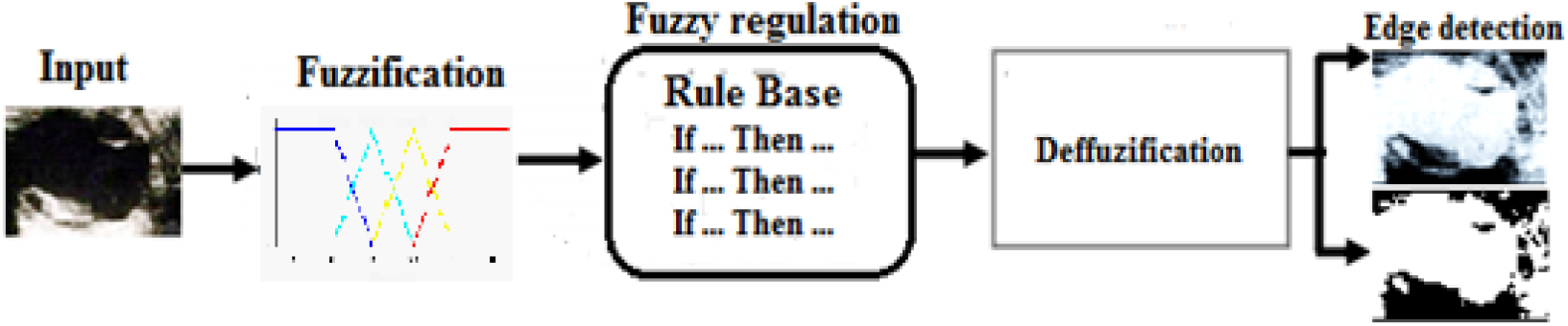
Fuzzy edge detection method

The steps utilized in pre-processing phases consisted of RGB to Gray Scale, noise elimination, contrast upgrading, and image processing. The ultrassonography and FP input images are transformed in gray scale image, using the equation IC = 0.333MR + 0.5MG + 0.1666MB, where IY indicates magnitude of tantamount gray quality image of RGB. The noise elimination was utilized the Hard Thresholding (HT) and Soft Thresholding (ST): 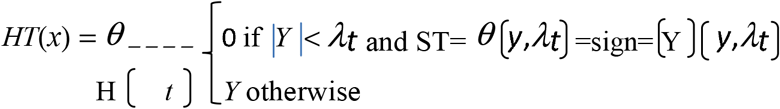

The *λ****t*** represents universal Threshold that was evaluated using the equation: 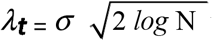 wherein *σ*indicates noise difference and N the dimension of image.

The contrast enhancement was calculated with the equations:

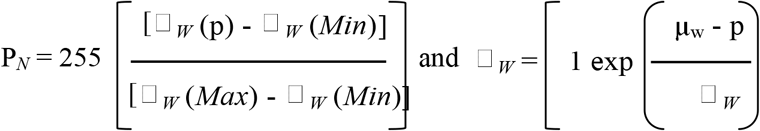

wherein min and max matches to minimum and maximum value of full image, μ_w_ is equivalent to local window mean, and □ *_W_* is equals local window standard deviation.

The image morphological processing implied equations for dilatation and erosion: Dilation: I_i_⊕A_1_={(p+q)/p∈ I_i_, q∈ A_1_} and Erosion: I_i_⊖A_1_={p∈ ^2^/(p+q) ∈I_i_}. In dilation and erosion image Ii is symbolized by the structural element Ai.

The Fuzzy-Calc (http://fuzzy-calc.appspot.com), an app to calculate fuzzy indicators was used with the objective to obtain a synthetic performance value for the study variables. The figure 2 demonstrates the simulation model of evaluation indexes using Fuzzy-Calc:

**Figure 2.**
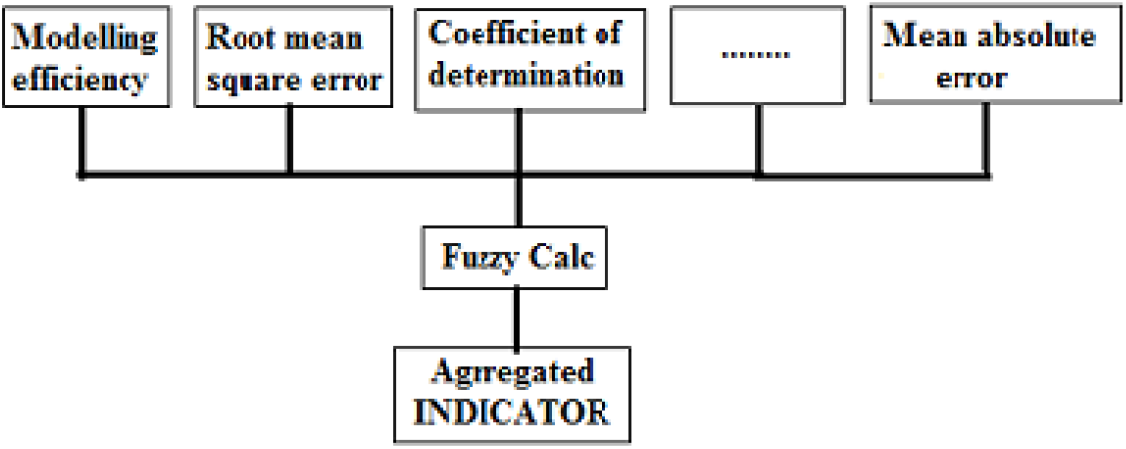
Fuzzy-Calc simulation model

The segmentation of according input variables in fuzzy logic system has been divided in two pattern: 1) the ultrasonography for determine to score of ultrasound ocular abnormalities (no retinal abnormality, vitreous hemorrhage, posterior vitreous detachment, epiretinal fibrosis, and retinal detachment), and, 2) the FP for determine to score of FP abnormalities (no retinal abnormality, four or more microaneurysms, four or more microaneurysms without hard or soft exudates, four or more microaneurysms with hard or soft exudates, and neovascularization).

The classification was suggested upon the estimated mean having a 95% confidence interval, based on the standard error of the mean from the Student’s t test. Three decisions were considered to define the classification: the presence of at least three classes (mild, moderate and severe), necessity of 95% confidence intervals superposition, and minimization of estimation errors^(7)^.

The value T is assigned for each retinal structure was calculated with follow equations: 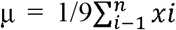 and 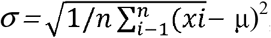, on what, N correspond total number of pixels, x_i_ indicate pixel values of full image.

## Results

We calculated the ratio of detected microaneurysms in FP to the total ultrasound image alterations. We categorize these images into mild group RD, moderately mild RD, moderate RD, moderately severe RD, severe RD, and very severe RD according to the percentage of the area damaged.

The ultrasonography images show the input no retinal abnormality, vitreous hemorrhage, posterior vitreous detachment, epiretinal fibrosis, and retinal detachment (Figure 3).

**Figure 3.**
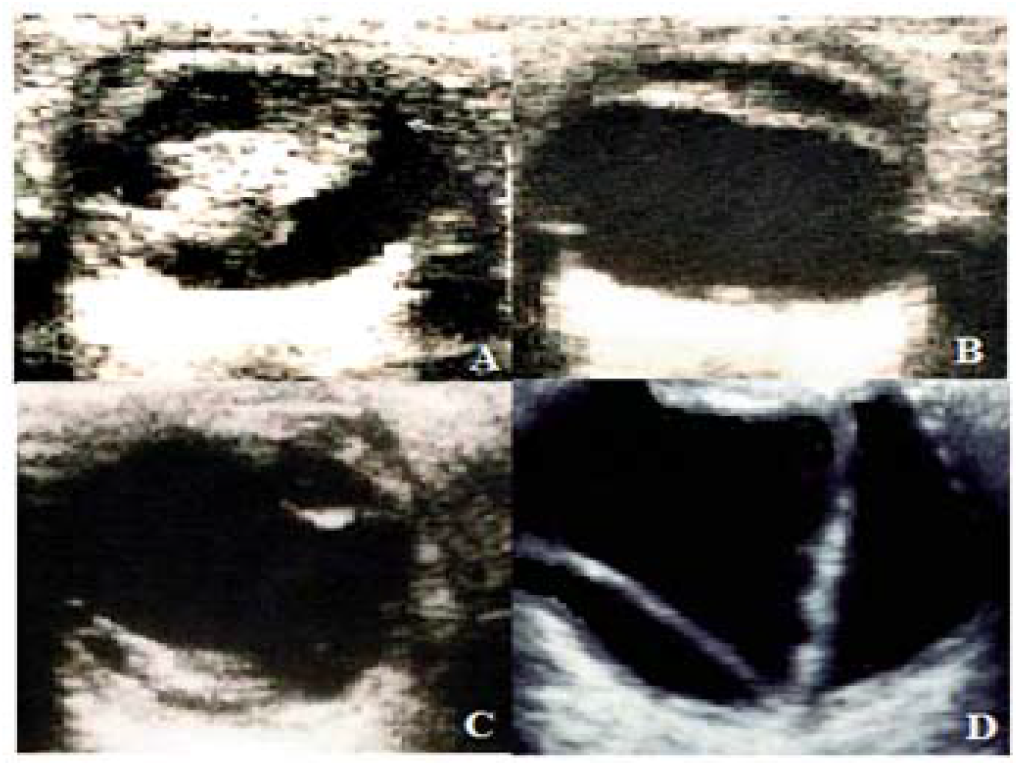
**A**: vitreous hemorrhage; B: posterior vitreous detachment; **D:** epiretinal fibrosis; **D:** retinal detachment **Source:** author’s collection

The FP images show the input no retinal abnormality, four or more microaneurysms, four or more microaneurysms without hard or soft exudates, four or more microaneurysms with hard or soft exudates, and neovascularization (Figure 4).

**Figure 4.**
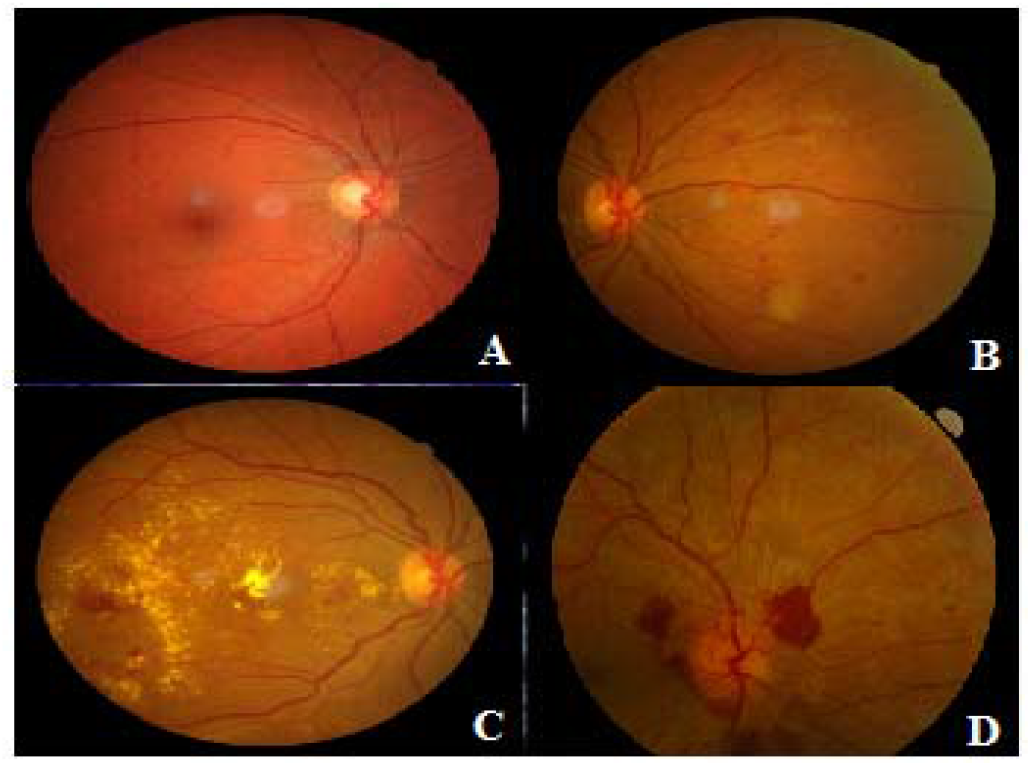
**A:** microsaneurysms; **B:** soft exudates; **C:** hard exudates; **D:** neovascularization **Source:** author’s collection.

Using MATLAB software (https://la.mathworks.com/products/matlab.html), automatic detection of secondary changes to DR is quickly performed. The images were pre-processed after a contrast improvement step that consisted of resizes the images to a uniform size of 500 × 500 pixels, morphological operators and then binarization. A total of 100 fuzzy rules extracted as a rule-base based on ophthalmologist and physician expert’s opinion. The FP images after pre-processing show the output no retinal abnormality, four or more microaneurysms without hard or soft exudates, four or more microaneurysms with hard or soft exudates, and neovascularization (Figure 5).

**Figure 5.**
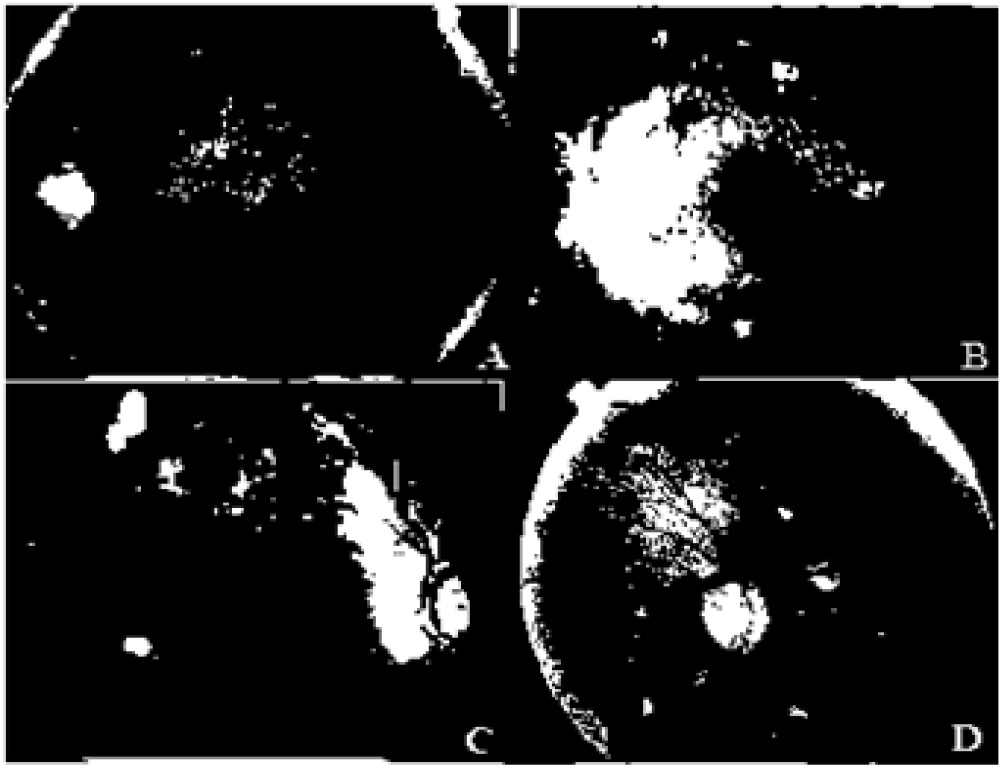
**A:** microaneurysms; **B:** soft exudates; **C:** hard exudates; **D:** neovascularization. **Source:** Research result

The table 1 shows the classification for DR suggested based on the 95% confidence interval of number of microaneurysms.

**Table 1.** Classification DR suggested based number of microaneurysms.

| Classification DR suggested | 95% confidence interval |
| --- | --- |
| Mild group | < 24.6 |
| Moderately mild | 24.6 - 48.0 |
| Moderate | 48.1 - 64.5 |
| Moderately severe | 64.6 - 77.0 |
| Severe | 77.1 - 92.7 |
| Very severe | > 92.7 |

We present in table 2 the results for the detection DR using fuzzy logic for six different ultrasound images. Therefore, the proposed DR severity classification considers the various 95% CI for the previously described classes, grounded on relationship with the found on ultrasound, as demonstrated in table 2. The results showed that is possible to develop fuzzy systems that can DR severity classification. Moreover, the results demonstrate that the fuzzy logic too can diagnose the various stages of DR.

**Table 2.**
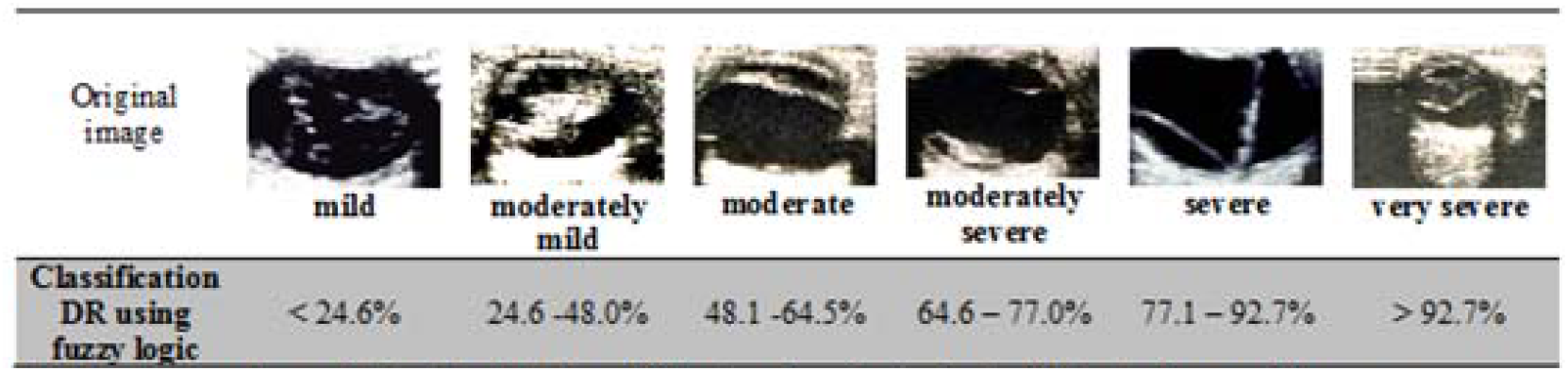
Shows the results for the detects on of DR using fuzzy logic for six different ultrasound images

|  |  |  |  |  |  |  |
| --- | --- | --- | --- | --- | --- | --- |
| Original image |  |  |  |  |  |  |
|  | mild | moderately mild | moderate | moderately severe | severe | very severe |
| Classification DR using fuzzy logic | < 24.6% | 24.6 - 48.0% | 48.1 - 64.5% | 64.6 - 77.0% | 77.1 - 92.7% | > 92.7% |

## Discussion

In this paper, we used the fuzzy logic with the severity classification proposal of DR, and results found demonstrated that the technique used can classify DR with elevated precision. Several scientists have introduced different fuzzy logic approaches for DR classification to raise confidence and decrease wrong classification secondary to human error, and language strategies has been implemented in decision-making with fuzzy interface^(8)^.

Ophthalmic ultrasonography has been used associated to associate with eye examination to complete the correct diagnosis in diabetic’s patients. Current evidence demonstrates the excellent performance of ocular ultrasound in evaluation of DR. Use of ophthalmic ultrasonography in patients with DR can provide essential information in detection and evaluation of DR complications^(9)^.

FP is one of the most practiced imaging procedures, owning a sensitivity and specificity quite superior to direct and indirect ophthalmoscopy. The FP is gold standard method for diagnosis of DR established by ETDRS group^(10)^. In 2016 Google company elaborated an algorithm to diagnose DR in retinal FP, and a study to evaluate this algorithm demonstrated a high specificity and sensitivity, with an area under the ROC curve of 0.99^(11)^.

The automation in evaluation of DR has been simplified by use of combining FP images and ocular ultrasonography using high resolution equipment, associated the processing digital and analysis of images through modern computational techniques. Thus, the diagnosis by computerized systems of the DR has been related by several authors^(12)^. In the present work we use morphological evaluations with FP and ultrasonography for fractionation and fuzzy logic for the identification process of features of DR. The FP aspects evaluated for diagnosis of DR were at least four or more microaneurysms with or without hard or soft exudates, and neovascularization, while following ultrasound findings were evaluated employing a semi-quantitative punctuation method, measuring the degree of vitreous hemorrhage, posterior vitreous detachment, epiretinal fibrosis, and retinal detachment.

Fuzzy logic was first described in 1965, it is a tool that produces a mechanism for reply to imprecise information based in qualitative language and transcribed to a numerical language (Boolean logic). Thus, fuzzy logic has been used indeed in medicine to diagnose the diseases based clinical and imaging data^(13,14)^. The fuzzy logic has been used in clinical research, especially in diabetes and your complications as DR, diabetic neuropathy, and diabetic nephropathy^(15,16)^. As already described above, we used the fuzzy logic with the severity classify objective of DR.

The fuzzy logic suggested in this study was based in four FP variables and four ultrasonography variables that went easy to do in routine eye exams. The analysis exposed here demonstrated that fuzzy logic is appropriate to classify the severity of DR likewise that retinal medical specialists.

The DR is a progressive microvascular complication of diabetes producing irreversible retinal deterioration. Thus, precocious diagnosis and preventive intervention are the best tools that may prevent or retard blindness, and consequently improve the quality of life of these individuals^(17)^. Taking into consideration the DR characteristics, we apply fuzzy logic to rank characteristics sonographic and FP related to DR. Posteriorly, a transformed fuzzy neural network was produced to improve the classification precision, and lastly we extract association rules between the chosen characteristics of DR to characterize their severity degree.

In the last years, several fuzzy system free and open source software were produced, through which simulations fuzzy logic can be performed quickly. In this work the evaluations were performed with Fuzzy-Calc, and the MATLAB package, the first an app was used calculate fuzzy indicators with the objective to obtain a synthetic performance value for the study variables, and MATLAB fuzzy logic toolbox was used for comparative study between FP and ultrasonography. The MATLAB software offer several tools and module used in this work was the Image Processing Toolbox^(18)^.

The medical diagnostic depend upon practice and expertise of medical practitioner. Fuzzy logic is a technique to establish whit accuracy what is imprecise in medicine, and has been playing a considerable role in the diagnosis and treatment of diseases^(19)^. Several researches employing models identification and image manipulation techniques to diagnose DR have been developed^(20)^. Our study as compared to the other studies was more embracing, because as it involved the use of FP and ultrasonography in different stages of the DR using fuzzy logic, and classifying the DR with high precision.

Thus, by the fuzzy logic, a DR classification was constructed supported on number of microaneurysms measurement with a simple practical application. Thus, the fuzzy logic technique use has been applied in many knowledge areas, and in medicine it has been used for diagnosis, diseases classification, and optimization of medical treatments.

## Data Availability

All data referred to in the manuscript are available for evaluation

## Disclosure of potential conflicts of interest

None of the authors have any potential conflicts of interest to disclose.

The manuscript was ethical approval of Ethics Committee of the Hospital de Base Luis Eduardo Magalhães - Itabuna - Bahia - Brazil.

## References

1. Yau JW, Rogers SL, Kawasaki R, Lamoureux EL, Kowalski JW, Bek T, et al. Global prevalence and major risk factors of diabetic retinopathy. Diabetes Care. 2012;35(3):556–64.

2. Bhagat N, Zarbin MA. (2019) Epidemiology, Risk Factors, and Pathophysiology of Diabetic Retinopathy. In: Bandello F, Zarbin M, Lattanzio R, Zucchiatti I. (eds) Clinical Strategies in the Management of Diabetic Retinopathy. Springer, Cham.

3. Wang W, Lo ACY. Diabetic Retinopathy: Pathophysiology and Treatments. Int J Mol Sci. 2018;19(6).

4. Andrade LJO, Bittencourt AMV, França CS. Sonographic ocular findings in diabetic retinopathy. Rev Ciênc Méd Biol. 2013;12(1):33–8.

5. Janghorbani A, Moradi MH. Fuzzy Evidential Network and Its Application as Medical Prognosis and Diagnosis Models. J Biomed Inform. 2017;72:96–107.

6. Grading diabetic retinopathy from stereoscopic color fundus photographs--an extension of the modified Airlie House classification. ETDRS report number 10. Early Treatment Diabetic Retinopathy Study Research Group. Ophthalmology. 1991;98(5 Suppl):786–806.

7. Meesad P, Yen GG. Accuracy, comprehensibility and completeness evaluation of a fuzzy expert system. IJUFKS. 2003;11:445–66.

8. Zhao J, Lin LY, Lin CM. A General Fuzzy Cerebellar Model Neural Network Multidimensional Classifier Using Intuitionistic Fuzzy Sets for Medical Identification. Comput Intell Neurosci. 2016;2016:8073279.

9. Zvomičanin J, Zvomičanin E. Use of ophthalmic B-scan ultrasonography in determining the causes of low vision in patients with diabetic retinopathy. Eur J Radiol Open. 2018;5:92.

10. Raman R, Srinivasan S, Virmani S, Sivaprasad S, Rao C, Rajalakshmi R. Fundus photograph-based deep learning algorithms in detecting diabetic retinopathy. Eye (Lond). 2019;33(1):97–109.

11. Gulshan V, Peng L, Coram M, Stumpe MC, Wu D, Narayanaswamy A, et al. Development and Validation of a Deep Learning Algorithm for Detection of Diabetic Retinopathy in Retinal Fundus Photographs. JAMA. 2016;316(22):2402–10.

12. Li F, Liu Z, Chen H, Jiang M, Zhang X, Wu Z. Automatic Detection of Diabetic Retinopathy in Retinal Fundus Photographs Based on Deep Learning Algorithm. Transl Vis Sci Technol. 2019;8(6):4.

13. Safdari R, Arpanahi HK, Langarizadeh M, Ghazisaiedi M, Dargahi H, Zendehdel K. Design a Fuzzy Rule-based Expert System to Aid Earlier Diagnosis of Gastric Cancer. Acta Inform Med. 2018;26(1):19–23.

14. Thukral S, Rana V. Versatility of fuzzy logic in chronic diseases: A review. Med Hypotheses. 2019;122:150–56.

15. Ibrahim S, Chowriappa P, Dua S, Acharya UR, Noronha K, Bhandary S, et al. Classification of diabetes maculopathy images using data-adaptive neuro-fuzzy inference classifier. Med Biol Eng Comput. 2015;53(12):1345–60.

16. Improta G, Mazzella V, Vecchione D, Santini S, Triassi M. Fuzzy logic-based clinical decision support system for the evaluation of renal function in post-Transplant Patients. J Eval Clin Pract. 2019 Nov 12.

17. Simó-Servat O, Hernández C, Simó R. Diabetic Retinopathy in the Context of Patients with Diabetes. Ophthalmic Res. 2019;62(4):211–17.

18. Gonzalez RC, Woods RE, Eddins SL. Digital image processing using MATLAB. Prentice Hall: Upper Saddle River, NJ, 2004.

19. Licata G. Probabilistic and fuzzy logic in clinical diagnosis. Intern Emerg Med. 2007 Jun;2(2):100–6.

20. Valverde C, Garcia M, Hornero R, Lopez-Galvez MI. Automated detection of diabetic retinopathy in retinal images. Indian J Ophthalmol. 2016 Jan;64(1):26–32.

